# Radiomic Profiling of Chest CT in a Cohort of Sarcoidosis Cases

**DOI:** 10.1101/2022.10.01.22280365

**Authors:** Nichole E Carlson, William Lippitt, Sarah M Ryan, Margaret Mroz, Briana Barkes, Shu-Yi Liao, Lisa A Maier, Tasha E Fingerlin

## Abstract

**Background:** High resolution computed tomography (HRCT) of the chest is increasingly used in clinical practice for sarcoidosis. Visual assessment of chest HRCTs in patients with sarcoidosis has high inter- and intra-rater variation. Radiomics offers a reproducible quantitative assessment of HRCT lung parenchyma and could be useful as an additional summary measure of disease. We develop radiomic profiles on HRCT and map them to radiologic, clinical, and patient reported outcomes.

**Research Question:** Can radiomic analysis of chest HRCT cluster patients into groups that are related to radiologic, clinical, and patient reported outcomes?

**Study Design and Methods:** Three-dimensional radiomic features were calculated on chest HRCT for both lungs from sarcoidosis cases enrolled in the Genomic Research in Alpha-1 Antitrypsin Deficiency and Sarcoidosis (GRADS) study (N=320). Robust and sparse K-means was used to cluster sarcoidosis cases using their radiomic profiles. Differences in patterns on visual assessment (VAS) by cluster were identified using chi-squared tests. Linear regression investigated how pulmonary function tests and patient reported outcomes differed between clusters with and without adjustment for other radiologic quantification.

**Results:** Radiomic-based clustering identified four clusters associated with both Scadding stage and Oberstein score (*P*<0.001). One of the clusters had markedly few abnormalities. Another cluster had consistently more abnormalities along with more Scadding stage IV. Average pulmonary function testing (PFT) differed between clusters, even after accounting for Scadding stage and Oberstein score (*P* <0.001), with one cluster having more obstructive disease. The most discriminative radiomic measures explained 10-15% of the variation in PFT beyond demographic variables. Shortness of breath, fatigue, and physical health differed by cluster (*P* <0.014).

**Interpretation:** Radiomic quantification of sarcoidosis identifies new subtypes representative of existing radiologic assessment and more predictive of pulmonary function. These findings provide evidence that radiomics may be useful for identifying new imaging-based disease phenotypes.

---

Sarcoidosis is a granulomatous interstitial lung disease which affects ∼ 110 thousand individuals in the United States^1^. Typical diagnosis is between 30-50 years of life, resulting in decreases in quality of life (QOL) and productivity^2^. Pulmonary disease occurs in over 90% of those with sarcoidosis^3^ with significant morbidity and mortality. Currently, visual assessment of chest radiography (CXR) is used to quantify lung abnormalities standardized via the Scadding staging^4^. Substantial variation exists in CT-based radiographic patterns within each Scadding stage limiting its utility in predicting prognosis even in the extreme stages^5^.

Chest high resolution computed tomography (HRCT) is increasingly used in clinical practice to monitor disease as it offers more detailed visualization of parenchymal abnormalities (PA) compared to CXR^4,6–8^. As with CXR, visual assessment of chest HRCT is used to evaluate abnormalities although there are limited standardized scoring metrics^4,9,10^. This is due in part to the diverse and heterogeneous patterns present on chest HRCT in sarcoidosis, often with multiple patterns noted. These complexities result in high inter- and intra-rater variation^11^. Recently, a Delphi study was undertaken to define phenotypes based on visual assessment, yet without assessment of clinical utility^12^. As not all patients have access to expert visual interpretation, more automated systems that quantify sarcoidosis chest HRCT could decrease variation and make HRCT more usable in routine clinical care.

Radiomics is when large numbers of quantitative features are extracted from medical images^13^. A radiomics panel computes summary measures of the distribution of the Hounsfield units (HU) along with summary measures of the spatial relationships of neighboring voxels^14^. The result is a characterization of image texture.

Radiomics have proved useful for quantifying HRCT in emphysema^7^, idiopathic pulmonary fibrosis^15,16^, interstitial lung disease^17,18^, diffuse lung disease^19^ and cancer^20^. Ryan et al.^21^ showed the potential utility of radiomics in sarcoidosis, comparing radiomic measures between sarcoidosis patients and controls. It remains unclear whether radiomics also has the potential to differentiate varied phenotypes *within* sarcoidosis patients and how radiomics relates to visual assessment (VAS). Radiomic profiles within patients with sarcoidosis that correlate with VAS, pulmonary function testing (PFT), and patient reported outcomes (PRO) would indicate that radiomics may serve as a useful refined measure to track change in the lung parenchyma over time than is possible with visual assessment.

The goal of this study was to develop a radiomic profile of chest HRCT in sarcoidosis using a phenotypically-diverse population of sarcoidosis participants from the Genomic Research in Alpha-1 Antitrypsin Deficiency and Sarcoidosis (GRADS) study^22^. We employ statistical clustering techniques and investigate the clinical utility of the clusters by quantifying their association with VAS, PFT and PRO^23,24^.

## Study Design and Methods

### Study design and participants

The sarcoidosis population was recruited in the multicenter NHLBI-funded GRADS study^22^ as cross-sectional observational cohort (N=368). This ancillary study has GRADS approval and all participants provided informed consent (IRB approval HS-2779 and HS-2780). More details of this cohort can be found in e-Appendix 1.

To comply with image biomarker standardization initiative (IBSI) recommendations^25^ details of the HRCT acquisition, processing and segmentation can be found in e-Appendix 1 and e-Tables 1 and 2. GRADS’ visual assessment score (VAS; Table 3, e-Appendix 1, e-Tables 3 and 4) was used to quantify overall Oberstein score^5,22^ along with additional information regarding presence of lymphadenopathy (LN), airway and vasculature distortion (AD), and PA (Table 3). Scadding stage was evaluated at the site using CXR (e-Appendix1).

**Table 1:**
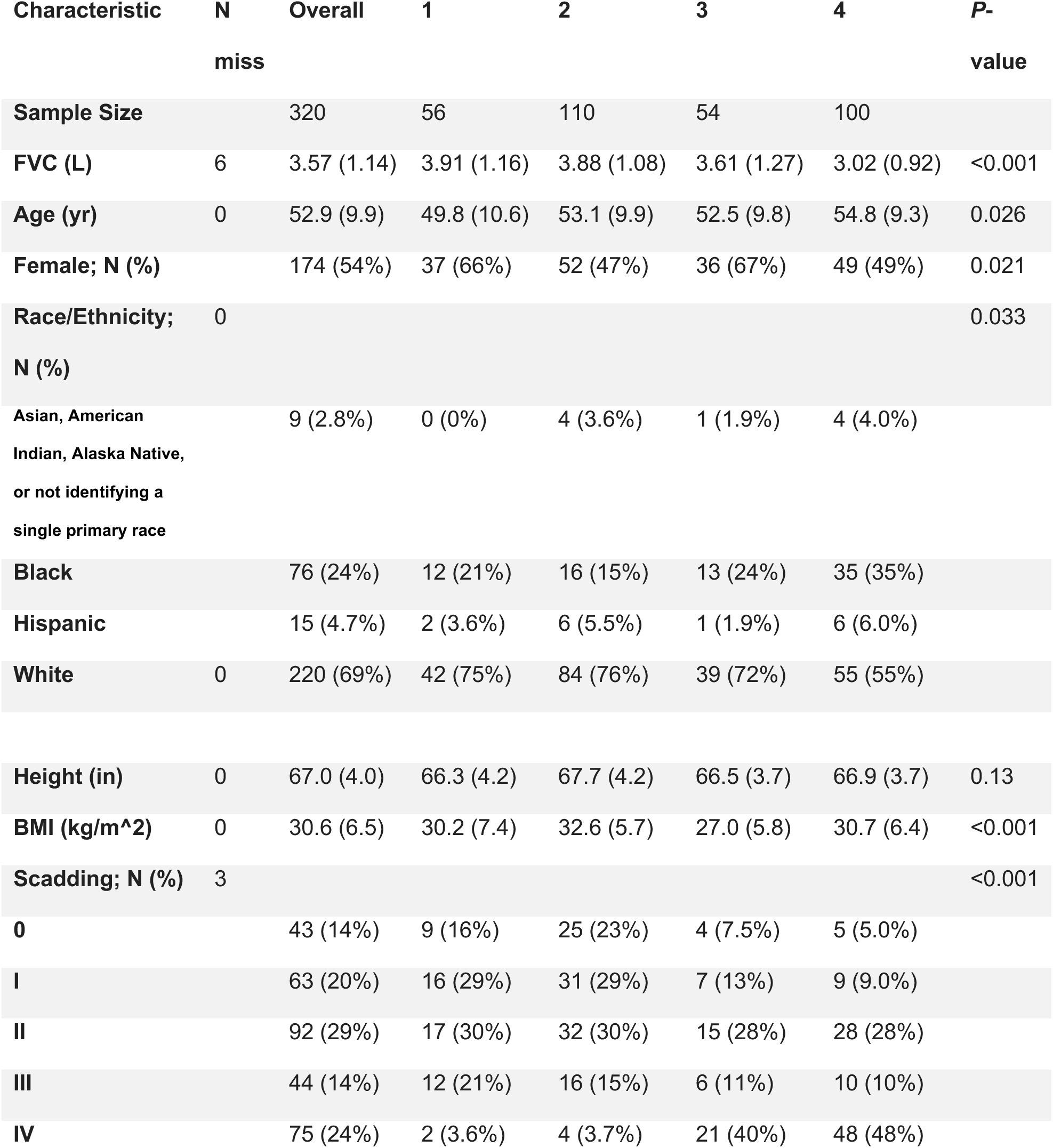
Patient demographics by radiomic cluster, ordered from least severe (1) to most severe (4) based on average FVC. Unless otherwise noted values are mean (SD).

PFT included pre-bronchodilator (pre-BD) forced expiratory volume at one second (FEV1), forced vital capacity (FVC), the ratio of FEV1 to FVC, and post-BD diffusing capacity of the lungs for carbon monoxide (DLCO). The PROs included the gastroesophageal reflux disease questionnaire (GERDQ)^26^, the University of California San Diego Shortness of Breath Questionnaire (SOBQ)^27^, two measures of fatigue (the Fatigue Assessment Scale [FAS]^28^ and Patient-Reported Outcomes Measurement Information System fatigue profile [PROMIS]^29^), the Cognitive Failure Questionnaire (CFQ)^30^, and the SF-12^31,32^ physical and mental subscales. The final analysis dataset included N=320 patients who had an analyzable HRCT and clinical data (e-Figure 1).

### Radiomic Analysis

We computed 44 first-order and 239 gray-level co-occurrence matrix (GLCM) radiomic features on each lung (566 features) using the lungct and RIA packages in R^33–36^.

These are open-source packages with published documentation and permit perfect reproducibility with transparent implementation, aligning with IBSI goals. Furthermore, this allowed a fully R based analysis pipeline and provided a more comprehensive radiomic panel. Radiomic features beyond those typically used in neuroscience and cancer and previously standardized by IBSI may be important in diffuse pulmonary disease. For consistency with package documentation, we use the nomenclature of the RIA package.

To calculate the gray level co-occurrence matrix (GLCM) features, the Hounsfield units from each HRCT were discretized into 16 bins with equal relative frequencies; then, the features were calculated in 13 directions, assuming a voxel distance of one; these features were summarized using the mean statistic ^34,35^.

The R ez.combat function harmonized radiomic measures across scanners^37–39^, while preserving the biological variability in age, height, BMI, sex, race-ethnicity and GRADS phenotype (e-Table 2).

Radiomic features were high-dimensional and repetitive (Figure 1). We used a decorrelation filter^40^ (e-Appendix 1) that prioritized first-order over second-order features to identify a representative feature subset for analysis. This reduced the features to 99. We used robust and sparse k-means^41^ to cluster participants (R package, RSKC). For outlier robustness, trimming was set at 0.1. The optimal bound on feature weights (9.5) and number of clusters (4) were simultaneously selected using a permutation approach and BCS-based Gap statistic^41^ after standardization^42^. Standardization was performed with respect to the permutation reference to permit comparability across bounds and cluster numbers. The top five discriminative features were selected for further investigation.

**Figure 1:**
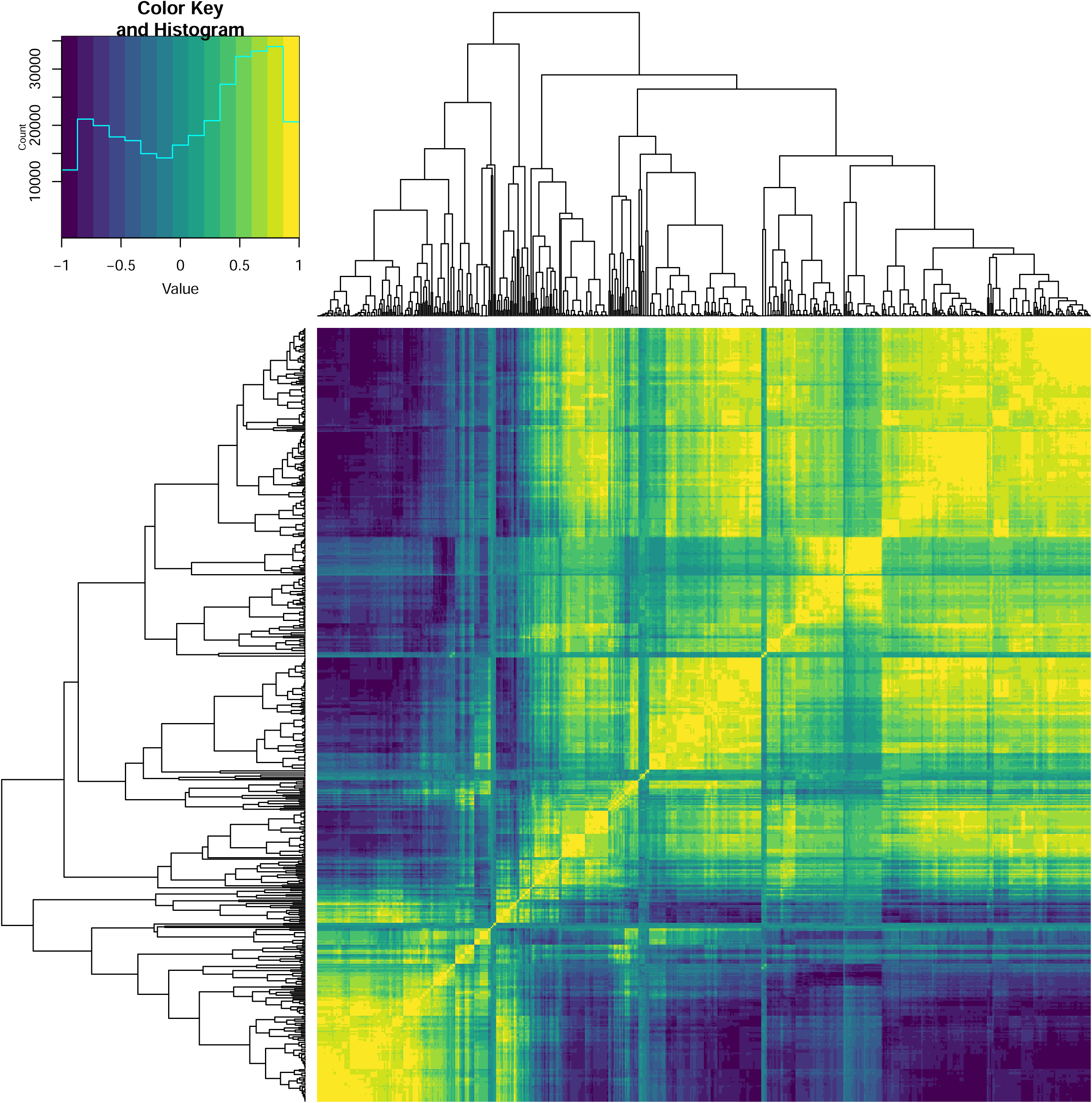
Heat map of the correlation between different radiomic measures for the entire population.

### Statistical Analysis and Validation

Descriptive statistics on VAS were computed by cluster and simulated Fisher’s exact tests used to assess cluster differences. Linear regression quantified associations between cluster, Scadding stage, and Oberstein Score, and each of the outcomes. We fitted univariable models with cluster, Scadding stage, or Oberstein score and then modeled them together to determine if cluster association remained significant accounting for other radiologic findings. Each outcome was modelled separately using a complete case analysis. Linear regression quantified associations between discriminative features and outcomes. All models were adjusted for age, sex, race/ethnicity (e-Appendix 1), height, and BMI. Analyses were conducted in R^36^.

This work was an unsupervised problem, which made traditional training and test validation approaches difficult given unknown true groupings. Instead, we conducted internal validation by applying our analysis pipeline under various conditions and with bootstrapped samples (e-Appendix 1).

## Results

Tables 1-2 shows the characteristics of the study population (N=320), 146 (46%) self-identified as male, 220 (68.8%) self-identified as non-Hispanic white, 76 (24%) as black, 15 (4.7%) as Hispanic, and 9 (2.8%) as one of Asian, American Indian, Alaska Native, not identifying a single primary race or missing (N=2). The average age was 53 (SD=10) years, average height 67.0 in (SD=4.0) and average BMI 30.6 kg/m^2^ (SD=6.5). By design participants spread across Scadding stages, with 43 (14%) stage 0, 63 (20%) stage 1, 92 (29%) stage 2, 44 (14%) stage 3 and 75 (24%) stage 4. The average FEV1 was 2.62 L (SD=0.89), FVC 3.57 L (SD=1.07), and DLCO 80.31 (SD=23.97). The population predominantly demonstrated non-obstructive FEV1/FVC ratio (N=235; 73%).

**Table 2:**
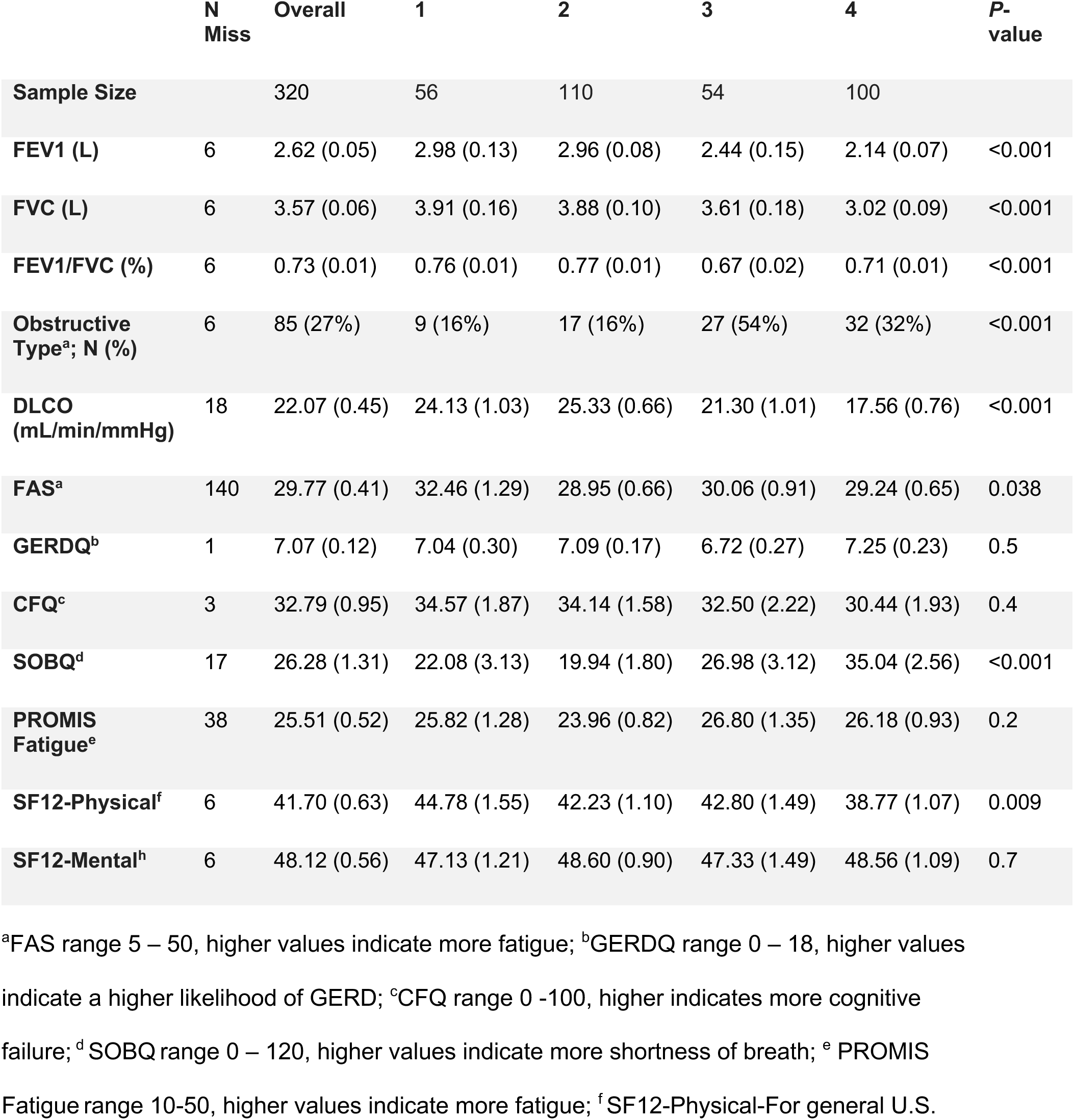
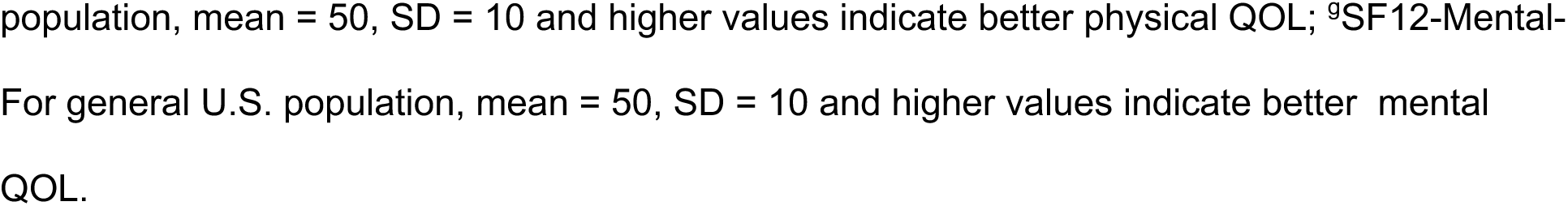
Summary measures of pulmonary function testing and self-reported outcomes by radiomic cluster ordered from least severe (1) to most severe (4) based on average FVC. Unless otherwise noted values are mean (SD).

|  | N<br>Miss | Overall | 1 | 2 | 3 | 4 | P-value |
| --- | --- | --- | --- | --- | --- | --- | --- |
| **Sample Size** |  | 320 | 56 | 110 | 54 | 100 |  |
| **FEV1 (L)** | 6 | 2.62 (0.05) | 2.98 (0.13) | 2.96 (0.08) | 2.44 (0.15) | 2.14 (0.07) | <0.001 |
| **FVC (L)** | 6 | 3.57 (0.06) | 3.91 (0.16) | 3.88 (0.10) | 3.61 (0.18) | 3.02 (0.09) | <0.001 |
| **FEV1/FVC (%)** | 6 | 0.73 (0.01) | 0.76 (0.01) | 0.77 (0.01) | 0.67 (0.02) | 0.71 (0.01) | <0.001 |
| **Obstructive Type<sup>a</sup>; N (%)** | 6 | 85 (27%) | 9 (16%) | 17 (16%) | 27 (54%) | 32 (32%) | <0.001 |
| **DLCO (mL/min/mmHg)** | 18 | 22.07 (0.45) | 24.13 (1.03) | 25.33 (0.66) | 21.30 (1.01) | 17.56 (0.76) | <0.001 |
| **FAS<sup>a</sup>** | 140 | 29.77 (0.41) | 32.46 (1.29) | 28.95 (0.66) | 30.06 (0.91) | 29.24 (0.65) | 0.038 |
| **GERDQ<sup>b</sup>** | 1 | 7.07 (0.12) | 7.04 (0.30) | 7.09 (0.17) | 6.72 (0.27) | 7.25 (0.23) | 0.5 |
| **CFQ<sup>c</sup>** | 3 | 32.79 (0.95) | 34.57 (1.87) | 34.14 (1.58) | 32.50 (2.22) | 30.44 (1.93) | 0.4 |
| **SOBQ<sup>d</sup>** | 17 | 26.28 (1.31) | 22.08 (3.13) | 19.94 (1.80) | 26.98 (3.12) | 35.04 (2.56) | <0.001 |
| **PROMIS Fatigue<sup>e</sup>** | 38 | 25.51 (0.52) | 25.82 (1.28) | 23.96 (0.82) | 26.80 (1.35) | 26.18 (0.93) | 0.2 |
| **SF12-Physical<sup>f</sup>** | 6 | 41.70 (0.63) | 44.78 (1.55) | 42.23 (1.10) | 42.80 (1.49) | 38.77 (1.07) | 0.009 |
| **SF12-Mental<sup>h</sup>** | 6 | 48.12 (0.56) | 47.13 (1.21) | 48.60 (0.90) | 47.33 (1.49) | 48.56 (1.09) | 0.7 |

<sup>a</sup>FAS range 5 – 50, higher values indicate more fatigue; <sup>b</sup>GERDQ range 0 – 18, higher values indicate a higher likelihood of GERD; <sup>c</sup>CFQ range 0 -100, higher indicates more cognitive failure; <sup>d</sup> SOBQ range 0 – 120, higher values indicate more shortness of breath; <sup>e</sup> PROMIS Fatigue range 10-50, higher values indicate more fatigue; <sup>f</sup> SF12-Physical-For general U.S.
population, mean = 50, SD = 10 and higher values indicate better physical QOL; <sup>9</sup>SF12-Mental- For general U.S. population, mean = 50, SD = 10 and higher values indicate better mental QOL.

**Table 3:** Distribution of VAS measures by radiomic cluster. Three participants were missing VAS and Scadding stage. All measures represent the abnormality was present (vs. absent) and are presented as N (%) unless otherwise noted. LN = lymphadenopathy; BVB = Bronchovascular Bundle.

| Characteristic | Overall | 1 | 2 | 3 | 4 | P-value |
| --- | --- | --- | --- | --- | --- | --- |
| **Sample Size** | **317** | **56** | **108** | **53** | **100** |  |
| **Mediastinal LN** | 172 (54%) | 19 (34%) | 60 (56%) | 29 (55%) | 64 (64%) | 0.003 |
| **Hilar LN** | 135 (43%) | 14 (25%) | 46 (43%) | 26 (49%) | 49 (49%) | 0.017 |
| **Micronodule** | 147 (46%) | 23 (41%) | 46 (43%) | 29 (55%) | 49 (49%) | 0.4 |
| **Airway and Vascular Distortion (AD)** |  |  |  |  |  |  |
| **BVB Distortion** | 202 (64%) | 19 (34%) | 53 (49%) | 43 (81%) | 87 (87%) | <0.001 |
| **Traction Bronchiectasis** | 129 (41%) | 6 (11%) | 25 (23%) | 32 (60%) | 66 (66%) | <0.001 |
| **Parenchymal Opacity and Distortion (PD)** |  |  |  |  |  |  |
| **Ground Glass** | 110 (35%) | 5 (8.9%) | 22 (20%) | 21 (40%) | 62 (62%) | <0.001 |
| **Honeycombing** | 25 (7.9%) | 0 (0%) | 2 (1.9%) | 4 (7.5%) | 19 (19%) | <0.001 |
| **Reticular Abnormality** | 88 (28%) | 3 (5.4%) | 17 (16%) | 21 (40%) | 47 (47%) | <0.001 |
| **Mosaic Attenuation** | 86 (27%) | 10 (18%) | 18 (17%) | 14 (26%) | 44 (44%) | <0.001 |
| **Interlobular Septal Thickening** | 54 (17%) | 5 (8.9%) | 14 (13%) | 7 (13%) | 28 (28%) | 0.007 |
| **Oberstein Overall; Mean (SE)** | 4.23<br>(0.18) | 2.11<br>(0.32) | 2.95<br>(0.26) | 5.32<br>(0.44) | 6.23<br>(0.28) | <0.001 |

### Radiomic Based Clustering

We identified four clusters (Table 1). Ordered from highest FVC to lowest, 56 (17.5%) patients were cluster 1, 110 (34.3%) cluster 2, 54 (16.9%) cluster 3, and 100 (31.3%) cluster 4. Cluster was associated with Scadding stage (*P*<0.001), but not a direct reflection of Scadding stage (Table 1). Descriptively, cluster 3 had the highest percentage of Scadding stage IV (48%). Clusters 1 and 2 each had approximately 60% of the patients with a Scadding stage of I or II, a higher proportion of stage 0, and limited Scadding stage IV present. Cluster 3 had the second highest observed Scadding stage IV (40%) with a similar percentage of Scadding stage II compared to clusters 1 and 2 (∼30%).

### Radiomic clustering and VAS

When descriptively investigating VAS patterns, the clusters reflected increased presence of VAS abnormalities with cluster 1 having low presence of nearly all airway and vascular distortions (AD) and PA and much lower average Oberstein scores than clusters 2-4 (Table 3; *P* <0.001). Cluster 2 reflected some increase in AD and PA with clusters 3 and 4 having a higher presence of AD and PA. For example, cluster 4 had at least double the presence of AD and PA compared to cluster 2. After adjustment for demographics, average Oberstein score for cluster 3 was 1.98 (SE=0.48) units and cluster 4 was 3.15 (SE=0.39) units higher than cluster 2 (p<0.001).

### Associations with PFT and PRO

After adjustment for demographics, average PFT differed between clusters (Figure 2; *P*<0.001). Clusters 2 and 3 had ∼0.3 L lower average FVC compared to clusters 1 and 4 had ∼0.9 L lower average FVC compared to cluster 1. Cluster 2 had ∼0.2 L lower average FVC compared to cluster 1 and cluster 3 had a larger decline with an average FEV1 ∼0.5 L lower than cluster 1. Cluster 4 had the lowest average FEV1 and ∼0.8 L lower than cluster 1. DLCO had a similar pattern to FVC. Cluster 3 had the most FEV1/FVC based obstruction (54%) with clusters 1 and 2 having the same amount of obstruction (16%) and cluster 4 falling in the middle (32%).

**Figure 2:**
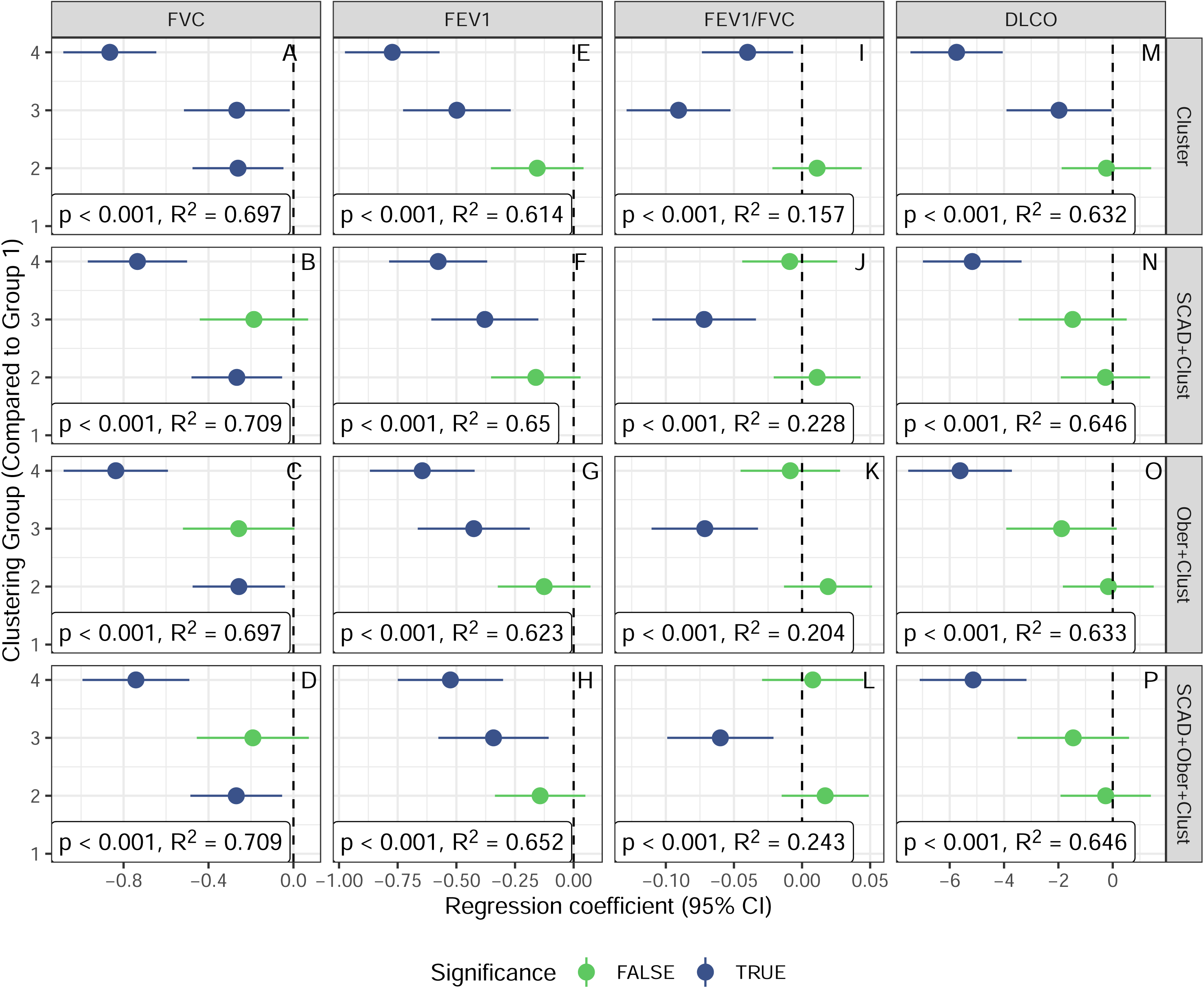
Radiomic cluster differences in average PFT values (columns) from regression models with adjustment for demographics (top row), then additionally adjusted for either Scadding stage or Oberstein score (middle two rows), then adjusted for both Scadding stage and Oberstein score (bottom row). The significance of the cluster differences from cluster 1 are in green (p>0.05) and blue (p<0.05). The bottom text in each panel is the overall p-value for the association between clustering and PFT outcome along with the R^2^.

After adjustment for demographics, PROs also differed between clusters but not consistently (Figure 3). Fatigue and Average shortness of breath (SOBQ) differed between clusters (P<0.014). Clusters 2 and 4 had a fatigue score that was more than 3 units lower on average compared to cluster 1 (p<0.016). Cluster 4 also had an average SOBQ ∼12 units higher than cluster 1 (p=0.003).

**Figure 3:**
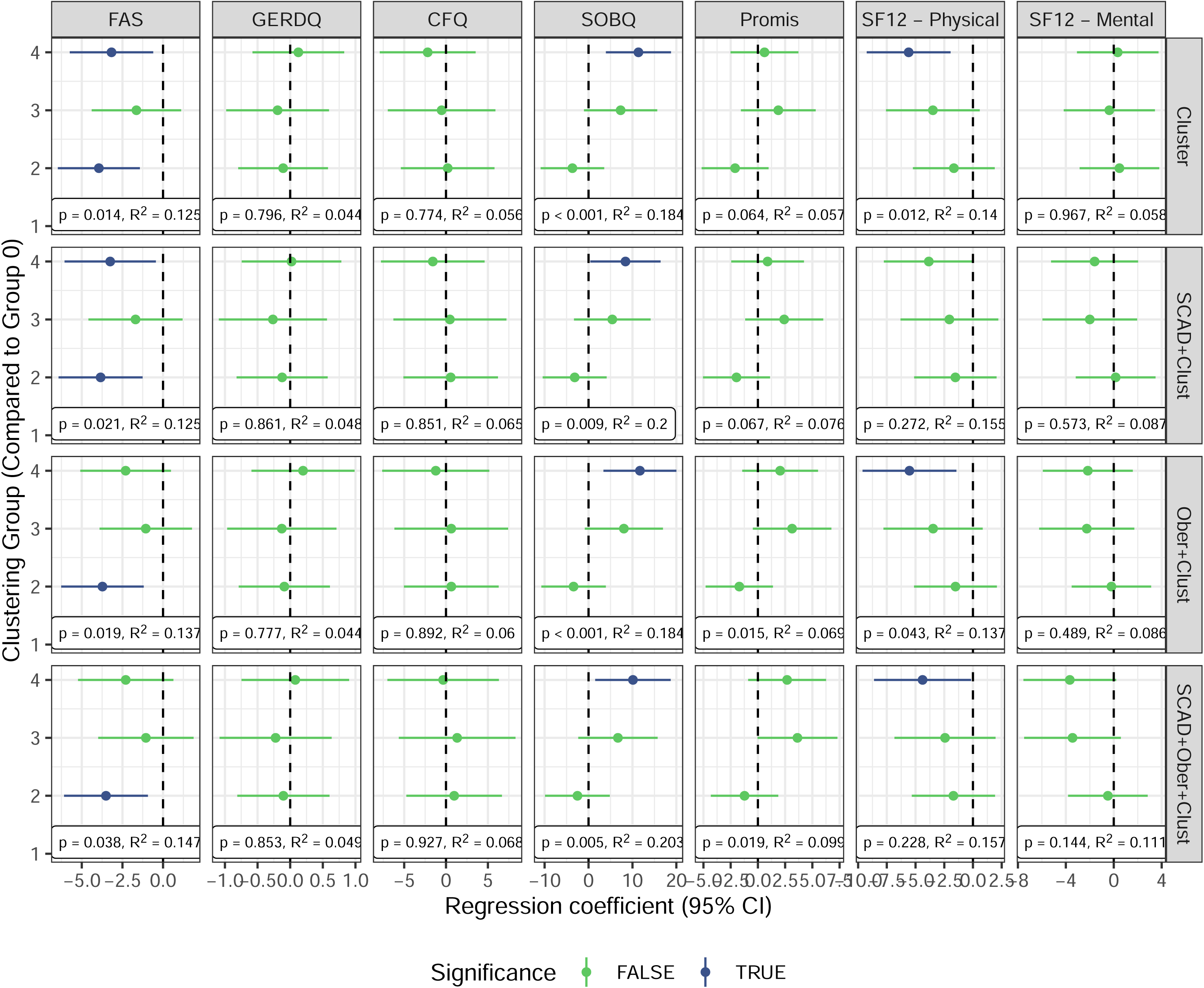
Radiomic cluster differences in average PRO values (columns) from regression models with adjustment for demographics (top row), then additionally adjusted for either Scadding stage of Oberstein score (middle two rows), then adjusted for both Scadding stage and Oberstein score (bottom row). The significance of the cluster differences from cluster 1 are in green (p>0.05) and blue (p<0.05). The bottom text in each panel is the overall p-value for the association between clustering and PFT outcome along with the R^2^.

The clusters remained significantly associated with PFT after adjusting for Scadding stage (Figure 2; *P*<0.001) or Oberstein score (Figure 2; *P*<0.001) and after adjusting for both simultaneously (Figure 2; *P*<0.001). Oberstein score was not significantly associated with FVC after adjustment for cluster (*P*=0.59) and became even less significant after further adjustment for Scadding stage (*P*=0.88). However, Scadding stage was significantly associated with FVC after adjustment for cluster and Oberstein score (*P*=0.019). The findings for Oberstein score with DLCO were like those of FVC. For FEV1, Oberstein score was still significantly associated after adjustment for cluster (*P*=0.011) but became insignificant after additional adjustment for Scadding stage (*P*=0.21). Scadding stage remained significant in all models (*P*<0.001). For FEV1/FVC, all three assessments (radiomics, Scadding, and Oberstein score) were significant (*P*<0.016).

Cluster also remained associated with fatigue and SOBQ score after adjustment for Scadding and Oberstein (*P*<0.038; Figure 3).

The five most discriminatory radiomic features included kurtosis, which is a measure of shape of the distribution of the HU from an image, as well as four summary measures from the gray level co-occurrence matrix (GLCM). The GLCM measure spatial correlation and similarity of the HU in image voxels near each other. Image visualization of cases with different values of kurtosis and two GLCM measures are shown in Figure 4 with distribution of HU in e-Figure 2 for maximum, median and minimum kurtosis levels in our population. The images visibly show the increase in observable PA (Figure 4) with lower kurtosis. In addition, Figure 5 shows how Oberstein Score separates based on kurtosis and GLCM Min. GLCM Min appears to be most related to severity with higher Oberstein score mapping with higher GLCM Min and Scadding stage is, in general, not associated with either measure.

**Figure 4:**
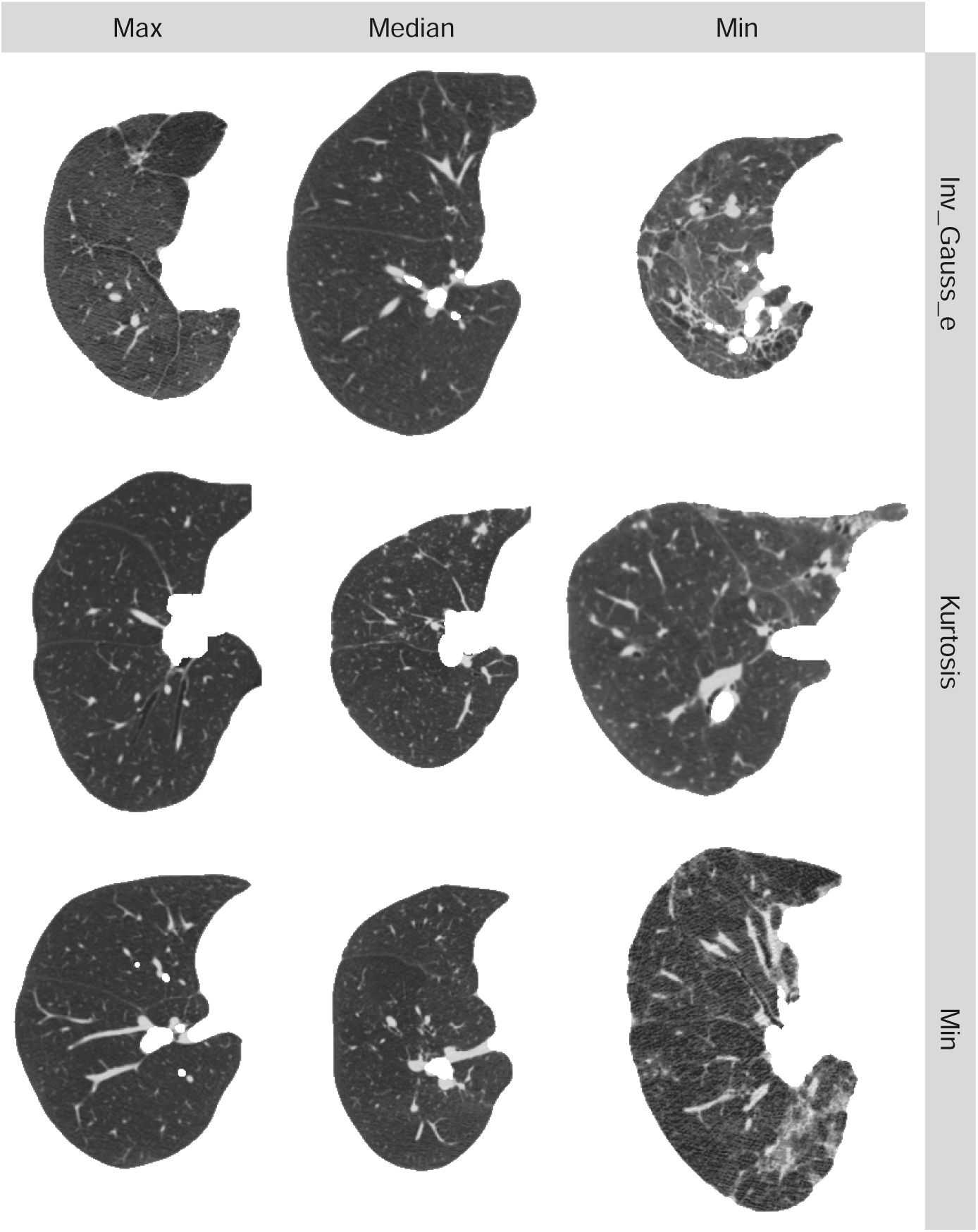
CT images in axial orientation for three patients with minimum, median, and maximum values for GLCM Gaussian (left column), the GLCM Inverse Gaussian, and the kurtosis.

**Figure 5:**
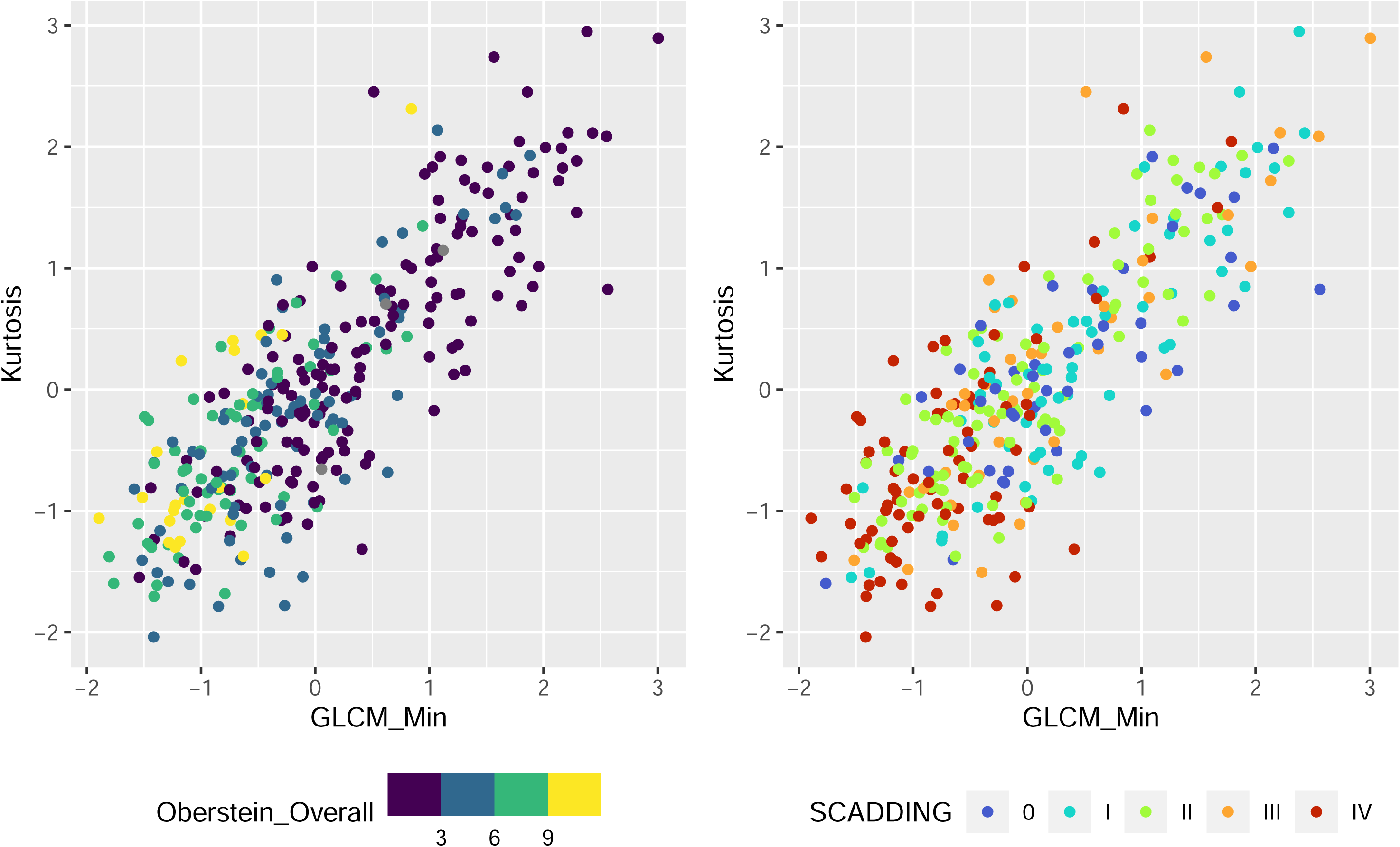
Separation of Oberstein score and Scadding based kurtosis (y-axis) and GLCM-Min (x-axis) two of the five most important radiomic variables in defining clusters. Colors represent the Oberstein score and Scadding stage.

The discriminatory radiomic measures were jointly associated with FVC, FEV1, FEV1/FVC and DLCO (*P*<0.001; Table 4 and e-Table 5 for PRO). The radiomic measures explained between 10 to 15% more variation in PFT than adjustment for demographics (age, race, sex, height and BMI) only. For comparison, Scadding stage explained between 4% and 12% more variation in PFT and Oberstein score explained between 2% and 8% more variation in PFT.

**Table 4:** Results of the multivariable regression analysis (coefficients and SE’s) of the five discriminatory radiomic measures for PFT.^d^.

|  | GLCM<br>Gaussian | GLCM Inv<br>Gaussian-E | Kurtosis | GLCM<br>Sum<br>Energy | GLCM Min | P-value | Rsqr | Rsqr -<br>base |
| --- | --- | --- | --- | --- | --- | --- | --- | --- |
| **FVC** | -0.02 (0.11) | -0.06 (0.08) | **0.44 (0.08)<sup>a</sup>** | 0.04 (0.09) | -0.06 (0.08) | <0.0001 | 0.726 | 0.622 |
| **FEV1** | -0.16 (0.10) | **-0.17 (0.08)<sup>c</sup>** | 0.08 (0.07) | 0.01 (0.08) | **0.23 (0.08)<sup>a</sup>** | <0.0001 | 0.619 | 0.514 |
| **FEV1/FVC** | **-0.04 (0.02)<sup>b</sup>** | **-0.03 (0.01)<sup>b</sup>** | **-0.07 (0.01)<sup>b</sup>** | -0.02 (0.01) | **0.09 (0.01)<sup>b</sup>** | <0.0001 | 0.199 | 0.049 |
| **DLCO** | **-2.23 (0.85)<sup>a</sup>** | **-1.65 (0.65)<sup>b</sup>** | **1.40 (0.62)<sup>c</sup>** | 0.50 (0.70) | -0.04 (0.68) | <0.0001 | 0.636 | 0.541 |

<sup>a</sup> P-value <0.001; <sup>b</sup> P-value<0.025; <sup>c</sup> P-value<0.035. <sup>d</sup>Each linear regression model included all five radiomic features and was additionally adjusted for age, gender, race/ethnicity, BMI and height. Bolded cells are statistically significant (p-values in the footnote). GLCM Gaussian is the sum of the GLCM with a Gaussian weight applied. GLCM-Inv Gaussian-E is the sum of the GLCM for the inverse Gaussian weighting scheme. GLCM Sum Energy is the sum of the squared values in the GLCM and a measure of uniformity. GLCM Min is the minimum of the GLCM. P-value is for the partial F-test of significance of any of the five radiomic measures.

**Table 5:**
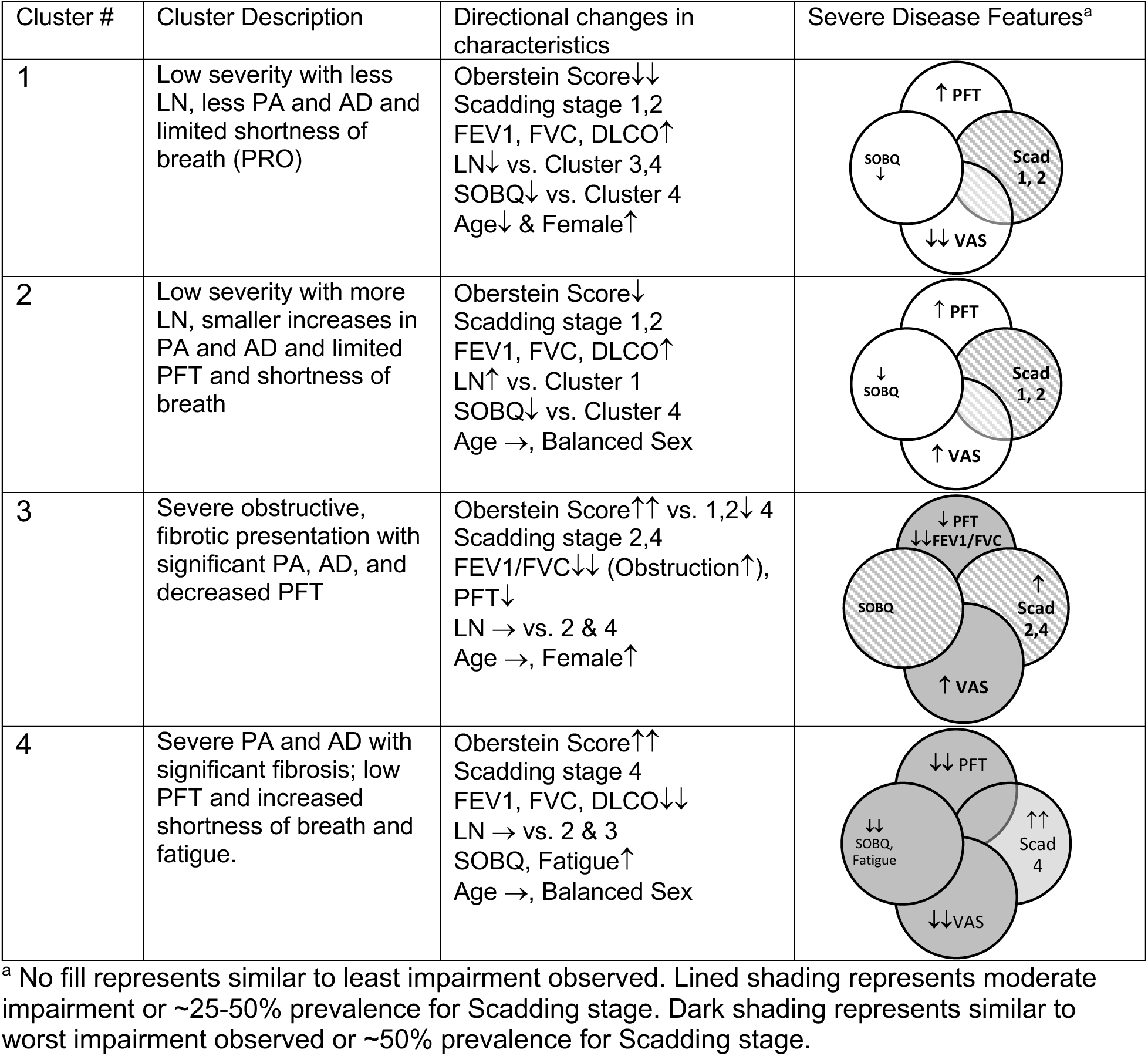
Holistic interpretation of radiomic clusters in terms of VAS, PFT, and PRO measures.

### Validation of Clustering

Detailed results can be found in e-Appendix 2, e-Figure 3. The validation analysis suggested only mild sensitivity of clustering to the analysis pipeline. There is more sensitivity to bootstrapped samples in how the sample is divided into clusters; however, cluster consistently remained a significant predictor of FVC across all sources of random perturbations in the pipeline. In all validation cases the proportions of *P*-values less than 0.01 were 100%.

## Discussion

Radiographic manifestations in sarcoidosis are protean. As a result, traditionally VAS is used for image characterization and not standardized. The utility of VAS in sarcoidosis is limited by the intra-observer and inter-observer variability. Radiomics is a more reproducible and computationally efficient approach to characterize HRCT. We used radiomics to characterize images from a large, phenotypically diverse cohort of sarcoidosis subjects and demonstrated that radiomics are associated with VAS, clinical and patient reported outcomes of disease. Using a common unsupervised learning approach^41^, we identified four radiomic based clusters. These clusters differed significantly by PFT, fatigue, and shortness of breath, two PROs. Notably, each cluster included a range of Scadding stages and cluster remained significantly associated with PFT after accounting for Scadding stage and Oberstein score. These data suggest that radiomics represents radiographic abnormalities that differ from Scadding stage and may be a better representation of image characterization to explain PFT and PRO.

A holistic assessment of the clusters showed two lower severity and two higher severity clusters (Table 5). One high severity cluster (4) had significant fibrosis, substantial presence of AD and PA on VAS, and decreased PFT, which resulted in more shortness of breath and fatigue. Cluster 3 represented those with obstructive spirometry and more presence of AD and PA than clusters 1 and 2, moderate fibrosis, and the lowest FEV1/FVC ratio. These interpretations have similarities to another recent attempt to subtype using clinical characteristics^43^. Unlike our work, that work had no radiomics or VAS. We both several low severity clusters with minimal PA. We both found an obstructive cluster and a severe fibrotic cluster. More recently, HRCT VAS phenotypes were proposed via a Delphi consensus study^12^. That work defined two major groups, those with fibrotic versus non-fibrotic findings, similar to our clusters with fibrotic (3 and 4) versus more non-fibrotic (1 and 2) findings. However, the VAS of their various subtypes of non-fibrotic and fibrotic CT findings were included in a number of our clusters. This may reflect the power in our sample size to define subtypes or possibly that these phenotypes or VAS overlap as noted in our study.

Demographic characteristics of the individual explained a sizable amount of the variation observed in PFT (R^2^=50-60%). Radiomics also explained a significant amount of additional variation in PFT (10-50%). This additional amount of variation explained is consistent with other quantitative imaging approaches such as CALIPER^17^ and those used for investigations of other lung conditions such as systemic sclerosis ^44^ and diffuse interstitial lung diseases ^45^.

Except in lung cancer research, much of the quantitative CT image analysis has focused on HU density^18,44^. We included summarization of both density and spatial characterization. More measures of GLCM appeared discriminative in the cluster analysis than densitometry measures. This implies that texture is perhaps more useful for differentiating PA in sarcoidosis.

Although it remains speculative as to why decreased kurtosis is associated with decreased FVC and DLCO, it is plausible that as kurtosis decreases, more severe PA are present that impact lung function. As kurtosis decreases, more severe PA are visual present that may impact the function of the lung (Figure 4). Severe PA in sarcoidosis are observed as higher HU values. In e-Figure we observed the presence of a higher frequency of higher HU values dampening the peak in the HU distribution and leading to a lower kurtosis. In addition, GLCM Min appears likely to measure disease severity based on the visual patterns with Oberstein in Figure 5.

The associations with cluster and PROs were less consistent, although important patterns emerged. Cluster 4 demonstrated decreased QOL (SF-12 physical) and worse shortness of breath (SOBQ), findings that are consistent with worse lung function and VAS scores in this cluster. Cluster explained substantially less variation in PROs (0 to 8%) compared to PFTs. While the tools we used to measure PROs are validated and used widely in sarcoidosis, they are not generally correlated with objective measures of lung function^46^. As an example, fatigue is a known multi-factorial PRO in sarcoidosis and may impact a number of other PRO^46,47^. Our finding that cluster is associated with decreased physical function and increased shortness of breath provides encouragement that more detailed characterization of lung abnormalities, like we developed here, could contribute to a better understanding of the current disconnect between measures of lung function and patient experiences.

This study had several strengths. To our knowledge, this was the largest cluster of sarcoidosis patients with research grade HRCT available allowing for a full 3-D based quantitative analysis. Radiomic and other quantitative imaging approaches have a major strength in that they can be computed in a time efficient and reliable, reproducible automated procedure. The radiomics package in R took only 3 minutes to compute all the 3-D radiomics measures. This means large quantities of scans can have radiomic profiles computed for aiding in decisions on what scans the radiologist might prioritize for further consideration for visual or clinician evaluation.

In addition to the high-quality and robust clinical data, the GRADS study also provides a comprehensive and consistently collected set of patient reported outcomes, which reflect important aspects of treatment decision-making processes^48^. The results internally validated suggest our approach is not over fitting these data.

This study is not without limitations. The GRADS study relied on enrollment from academic centers and could be skewed to a population that was referred for worse disease or was near one of the centers, which were primarily localized to the Eastern US. The demographic is predominantly white and of higher SES as a result and thus may not provide full representation of the spectrum of disease. In addition, many subjects had disease for a decade or longer and were treated. While the same protocol was used across study sites for obtaining the CT images we studied, the scanners themselves differed. Harmonization was performed to mitigate systematic differences due to scanner type. In addition, the distribution of PFT measures is not dependent on scanner type. As such, we expect any differences in the radiomic measures due to scanner type to be non-differential as they relate to PFTs. Finally, the number of clusters we identified may not directly translate to other populations of sarcoidosis or be the unique optimal solution. There are multiple ways to choose the optimal number of clusters. We used an alternative radiomics package with a broader set of measures. However, not all these measures were part of IBSI work. We used open-source software accessible to any reader for verification of our findings and to conduct comparisons with other software.

### Interpretation

In summary, this work provides evidence that in sarcoidosis radiomic quantification is useful for classifying abnormality of the lung along with pulmonary function and to a lesser degree patient reported outcomes. Future work should evaluate the potential of radiomics to capture small changes over time not easily detected by other assessment to further evaluate the potential of radiomics serving as a clinically useful quantitative measure of sarcoidosis presentation and progression.

## Supplementary Material Statement

This article has an online supplement, which is accessible from this issue’s table of contents online.

## Summary of COI

NEC, WLL, MM, TEF, SL, BB, TEF have received grants from the National Institutes of Health (R01 HL114587; R01 HL142049; U01 HL112695; U01 HL112707; U01 HL112707, U01 HL112694, U01 HL112695, U01 HL112696, U01 HL112702, U01 HL112708, U01 HL112711, U01 HL112712)

WLL is additionally support on National Institutes of Health 5T32HL007085

LAM has received grants from the National Institute of Health (R01HL140357, R01HL142049, R01 HL114587; U01 HL112695; U01 HL112707; U01 HL112707, U01 HL112694, U01 HL112695, U01 HL112696, U01 HL112702, U01 HL112708, U01 HL112711, U01 HL112712, and R01HL136681), Ann Theodore Foundation, the FSR, Mallinckrodt Pharmaceuticals, and the University of Cincinnati (Mallinckrodt Pharmaceuticals Foundation Grant) and serves on the Scientific Advisory Board for FSR, and Boeringer Ingelheim.

### Funding

This work is supported by NIH grants: R01 HL114587; R01 HL142049; U01 HL112695; U01 HL112707; U01 HL112707, U01 HL112694, U01 HL112695, U01 HL112696, U01 HL112702, U01 HL112708, U01 HL112711, U01 HL112712 and 5T32HL007085

## Data Availability

Data is available via the process laid out by the GRADS consortium.

## Acknowledgements and Author contributions

NEC is responsible for all content in the manuscript.

N.E.C. Substantial contributions to the conception or design of the work, lead biostatistical analysis and interpretation of the data and wrote manuscript. W.L. developed the filtering method for this analysis, performed the sensitivity and validation analyses and edited the manuscript. S.M.R. performed image processing, conducted statistical analysis, prepared figures and table, interpreted the data, and contributed to the manuscript. M.M. coordinated the study, cleaned data, and contributed to the manuscript. B.B. supported data acquisition, data cleaning, project management, and contributed to the manuscript. S.L. contributed to the scientific rationale and interpretation of the data and contributed to the manuscript. L.A.M. Substantial contributions to the conception or design of the work, interpretation of the data and wrote the manuscript. T.E.F. Substantial contributions to the conception or design of the work, interpretation of the data and wrote the manuscript.

This work was supported by the National Institutes of Health (R01 HL114587; R01 HL142049; U01 HL112695; T32HL007085). Data from the GRADS study was used, which was funded by the NIH grant U01 HL112707 entitled “Sarcoidosis and A1AT Genomics and Informatics Centre”, as well as others (U01 HL112707, U01 HL112694, U01 HL112695, U01 HL112696, U01 HL112702, U01 HL112708, U01 HL112711, U01 HL112712). The content is solely the responsibility of the authors and does not necessarily represent the official views of the National Heart, Lung, and Blood Institute or the National Institutes of Health.

## Abbreviations

AD: Airway and Vascular Distortion
BMI: body mass index
BVB: bronchovascular bundle
HRCT: High resolution chest computed tomography
CXR: chest X-ray
DLCO: diffusing capacity for carbon monoxide
FEV1: forced expiratory volume in one second
FEV1/FVC: ratio of FEV1 to FVC
FVC: forced vital capacity
GIC: Genetics and Informatics Core
GLCM: Gray level co-occurrence matrix
GRADS: Genomic Research in Alpha-1 Anti-trypsin Deficiency and Sarcoidosis
IRB: institutional review board
LN: lymphadenopathy
PA: parenchymal abnormalities
PFT: Pulmonary Function Test
QOL: Quality of life
VAS: visual assessment score

## Take-Home Points

Study Question: Is radiomics a useful quantitative approach for defining radiographic subtypes in sarcoidosis?

Results: Radiomics find four radiographic subtypes in sarcoidosis that are related to visual assessment and more predictive of clinical measures such as pulmonary function that visual assessment.

Interpretation: Radiomics analysis suggests four subtypes of sarcoidosis ranging from low severity with limited radiographic abnormality and normal pulmonary function to severe with significant abnormality on chest CT and either obstruction based on pulmonary function or reduced pulmonary function with fibrotic presentation on image.

## References

1. Erdal BS, Clymer BD, Yildiz VO, Julian MW, Crouser ED. Unexpectedly high prevalence of sarcoidosis in a representative U.S. Metropolitan population. Respir Med 2012;106(6):893–899.

2. Cox CE, Donohue JF, Brown CD, Kataria YP, Judson MA. The Sarcoidosis Health Questionnaire: a new measure of health-related quality of life. Am J Respir Crit Care Med 2003;168(3):323–329.

3. Wasfi YS, Rose CS, Murphy JR, et al. A new tool to assess sarcoidosis severity. Chest 2006;129(5):1234–1245.

4. Scadding JG. Prognosis of intrathoracic sarcoidosis in England. A review of 136 cases after five years’ observation. Br Med J 1961;2(5261):1165–1172.

5. Oberstein A, Zitzewitz H von, Schweden F, Müller-Quernheim J. Non invasive evaluation of the inflammatory activity in sarcoidosis with high-resolution computed tomography. Sarcoidosis Vasc Diffuse Lung Dis Off J WASOG 1997;14(1):65–72.

6. Drent M, De Vries J, Lenters M, et al. Sarcoidosis: assessment of disease severity using HRCT. Eur Radiol 2003;13(11):2462–2471.

7. Sluimer I, Schilham A, Prokop M, Ginneken B van. Computer analysis of computed tomography scans of the lung: a survey. IEEE Trans Med Imaging 2006;25(4):385– 405.

8. Keijsers RGM, Heuvel DAF van den, Grutters JC. Imaging the inflammatory activity of sarcoidosis. Eur Respir J 2013;41(3):743–751.

9. Moller DR. Negative clinical trials in sarcoidosis: failed therapies or flawed study design? Eur Respir J 2014;44(5):1123–1126.

10. Nunes H, Uzunhan Y, Gille T, Lamberto C, Valeyre D, Brillet P-Y. Imaging of sarcoidosis of the airways and lung parenchyma and correlation with lung function. Eur Respir J 2012;40(3):750–765.

11. Van den Heuvel DA, Jong PA de, Zanen P, et al. Chest Computed Tomography-Based Scoring of Thoracic Sarcoidosis: Inter-rater Reliability of CT Abnormalities. Eur Radiol 2015;25(9):2558–2566.

12. Desai SR, Sivarasan N, Johannson KA, et al. High-resolution CT phenotypes in pulmonary sarcoidosis: a multinational Delphi consensus study. Lancet Respir Med [Internet] 2023 [cited 2024 Jan 9];0(0). Available from: https://www.thelancet.com/journals/lanres/article/PIIS2213-2600(23)00267-9/fulltext

13. Kumar V, Gu Y, Basu S, et al. Radiomics: the process and the challenges. Magn Reson Imaging 2012;30(9):1234–1248.

14. Haralick RM. Statistical and structural approaches to texture. Proc IEEE 1979;67(5):786–804.

15. Park YS, Seo JB, Kim N, et al. Texture-based quantification of pulmonary emphysema on high-resolution computed tomography: comparison with density-based quantification and correlation with pulmonary function test. Invest Radiol 2008;43(6):395–402.

16. Humphries SM, Yagihashi K, Huckleberry J, et al. Idiopathic Pulmonary Fibrosis: Data-driven Textural Analysis of Extent of Fibrosis at Baseline and 15-Month Follow-up. Radiology 2017;285(1):270–278.

17. Ungprasert P, Wilton KM, Ernste FC, et al. Novel Assessment of Interstitial Lung Disease Using the “Computer-Aided Lung Informatics for Pathology Evaluation and Rating” (CALIPER) Software System in Idiopathic Inflammatory Myopathies. Lung 2017;195(5):545–552.

18. Ash SY, Harmouche R, Vallejo DLL, et al. Densitometric and local histogram based analysis of computed tomography images in patients with idiopathic pulmonary fibrosis. Respir Res [Internet] 2017 [cited 2018 Jan 8];18. Available from: https://www.ncbi.nlm.nih.gov/pmc/articles/PMC5340000/

19. Wang J, Li F, Doi K, Li Q. Computerized detection of diffuse lung disease in MDCT: the usefulness of statistical texture features. Phys Med Biol 2009;54(22):6881– 6899.

20. Lee G, Lee HY, Park H, et al. Radiomics and its emerging role in lung cancer research, imaging biomarkers and clinical management: State of the art. Eur J Radiol 2017;86:297–307.

21. Ryan SM, Fingerlin TE, Mroz M, et al. Radiomic measures from chest high-resolution computed tomography associated with lung function in sarcoidosis. Eur Respir J 2019;54(2).

22. Moller DR, Koth LL, Maier LA, et al. Rationale and Design of the Genomic Research in Alpha-1 Antitrypsin Deficiency and Sarcoidosis (GRADS) Study. Sarcoidosis Protocol. Ann Am Thorac Soc 2015;12(10):1561–1571.

23. Michielsen HJ, De Vries J, Van Heck GL, Van de Vijver FJR, Sijtsma K. Examination of the Dimensionality of Fatigue: The Construction of the Fatigue Assessment Scale (FAS). Eur J Psychol Assess 2004;20(1):39–48.

24. Cella D, Riley W, Stone A, et al. The Patient-Reported Outcomes Measurement Information System (PROMIS) developed and tested its first wave of adult self-reported health outcome item banks: 2005-2008. J Clin Epidemiol 2010;63(11):1179–1194.

25. Zwanenburg A, Vallières M, Abdalah MA, et al. The Image Biomarker Standardization Initiative: Standardized Quantitative Radiomics for High-Throughput Image-based Phenotyping. Radiology 2020;295(2):328–338.

26. Jones R, Junghard O, Dent J, et al. Development of the GerdQ, a tool for the diagnosis and management of gastro-oesophageal reflux disease in primary care. Aliment Pharmacol Ther 2009;30(10):1030–1038.

27. Eakin EG, Resnikoff PM, Prewitt LM, Ries AL, Kaplan RM. Validation of a New Dyspnea Measure: The UCSD Shortness of Breath Questionnaire. Chest 1998;113(3):619–624.

28. Michielsen HJ, De Vries J, Van Heck GL, Van de Vijver FJR, Sijtsma K. Examination of the Dimensionality of Fatigue: The Construction of the Fatigue Assessment Scale (FAS). Eur J Psychol Assess 2004;20(1):39–48.

29. Cella D, Riley W, Stone A, et al. The Patient-Reported Outcomes Measurement Information System (PROMIS) developed and tested its first wave of adult self-reported health outcome item banks: 2005-2008. J Clin Epidemiol 2010;63(11):1179–1194.

30. Broadbent DE, Cooper PF, FitzGerald P, Parkes KR. The Cognitive Failures Questionnaire (CFQ) and its correlates. Br J Clin Psychol 1982;21(1):1–16.

31. Ware JE, Kosinski M, Keller SD. A 12-Item Short-Form Health Survey: Construction of Scales and Preliminary Tests of Reliability and Validity. Med Care 1996;34(3):220–233.

32. Ware J, Kosinski M, Turner-Bowker D, Gandek B. How to score SF-12 items. SF-12 V2 Score Version 2 SF-12 Health Surv 2002;29–38.

33. Ryan SM, Vestal B, Maier LA, Carlson NE, Muschelli J. Template Creation for High-Resolution Computed Tomography Scans of the Lung in R Software. Acad Radiol 2020;27(8):e204–e215.

34. Kolossváry M, Kellermayer M, Merkely B, Maurovich-Horvat P. Cardiac Computed Tomography Radiomics: A Comprehensive Review on Radiomic Techniques. J Thorac Imaging 2018;33(1):26–34.

35. Kolossváry M, Karády J, Szilveszter B, et al. Radiomic Features Are Superior to Conventional Quantitative Computed Tomographic Metrics to Identify Coronary Plaques With Napkin-Ring Sign. Circ Cardiovasc Imaging 2017;10(12):e006843.

36. R Core Team. R: A Language and Environment for Statistical Computing [Internet]. Vienna, Austria: R Foundation for Statistical Computing; 2018. Available from: https://www.R-project.org/

37. Fortin J-P, Parker D, Tunç B, et al. Harmonization of multi-site diffusion tensor imaging data. NeuroImage 2017;161:149–170.

38. Fortin J-P, Cullen N, Sheline YI, et al. Harmonization of cortical thickness measurements across scanners and sites. NeuroImage 2018;167:104–120.

39. Johnson WE, Li C, Rabinovic A. Adjusting batch effects in microarray expression data using empirical Bayes methods. Biostat Oxf Engl 2007;8(1):118–127.

40. Lippit WL. Clustering with Highly Correlated Features. MS Thesis Univ Colo 2022;

41. Kondo Y, Salibian-Barrera M, Zamar R. RSKC : An R Package for a Robust and Sparse K-Means Clustering Algorithm. J Stat Softw [Internet] 2016 [cited 2020 May 27];72(5). Available from: http://www.jstatsoft.org/v72/i05/

42. Witten DM, Tibshirani R. A framework for feature selection in clustering. J Am Stat Assoc 2010;105(490):713–726.

43. Lin NW, Arbet J, Mroz MM, et al. Clinical phenotyping in sarcoidosis using cluster analysis. Respir Res 2022;23(1):88.

44. Camiciottoli G, Orlandi I, Bartolucci M, et al. Lung CT Densitometry in Systemic Sclerosis: Correlation With Lung Function, Exercise Testing, and Quality of Life. Chest 2007;131(3):672–681.

45. Shin KE, Chung MJ, Jung MP, Choe BK, Lee KS. Quantitative Computed Tomographic Indexes in Diffuse Interstitial Lung Disease: Correlation With Physiologic Tests and Computed Tomography Visual Scores. J Comput Assist Tomogr 2011;35(2):266–271.

46. Thunold RF, Løkke A, Cohen AL, Hilberg O, Bendstrup E. Patient Reported Outcome Measures (PROMs) in Sarcoidosis. Sarcoidosis Vasc Diffuse Lung Dis 2017;34(1):2–17.

47. Kampstra NA, Grutters JC, Beek FT van, et al. First patient-centred set of outcomes for pulmonary sarcoidosis: a multicentre initiative. BMJ Open Respir Res 2019;6(1):e000394.

48. Wijsenbeek MS, Culver DA. Treatment of Sarcoidosis. Clin Chest Med 2015;36(4):751–767.

