## Supplemental Materials for "Radiomic Profiling of Chest CT in a Cohort of Sarcoidosis Cases"

**Supplementary Material** "Radiomic Profiling of Chest CT in a Cohort of Sarcoidosis Cases"

### E-Appendices, Tables, and Figures

#### E-Appendix 1: Supplementary Methods Details

*GRADS description*

In brief, eligible participants were between the ages of 18 and 85 (N=368). Participants were enrolled into nine pre-specified clinically-described phenotypes to ensure a spectrum of disease manifestations with a focus on pulmonary disease manifested by Scadding stage, treatment, and other specific organ involvement. To be included in the GRADS cohort, the subject had to (a) have a diagnosis of sarcoidosis established by consensus criteria (ATS/ERS)[1] with confirmation by either biopsy or manifestations consistent with acute sarcoidosis (Löfgren's syndrome) in the absence of other known diagnosis.

Spirometry included forced expiratory volume in one second (FEV1), forced vital capacity (FVC), and FEV1 to FVC ratio (FEV1/FVC), and single-breath carbon monoxide diffusing capacity (DLCO). These measures were obtained for all subjects and DLCO was corrected for site elevation according to established criteria[2,3].

*Measurement of Self-Identified Race and Ethnicity and Treatment in Analysis*

A combined category for self-reported primary race and ethnicity was constructed from available data due to modest numbers in several categories. Non-Hispanic participants identifying as primarily white or black were classified as white or black participants respectively. Hispanic participants were indicated as such regardless of primary race identification (of which there were very few that indicated Hispanic ethnicity and a non-white race). Participants identifying as non-Hispanic and identifying primarily as Asian, American Indian, or Alaska Native, or not identifying a single primary race (identifying as multi-racial, having no primary race, or having unknown primary race) were combined into a single category for Table 1 summary measures and statistical analysis to address low event rates across groups and reduce accidental identification. Combined race/ethnicity was further collapsed to a binary variable of non-Hispanic white vs. not non-Hispanic white for statistical analysis given low events in radiomic groups. This variable was used as an adjustment factor in all adjusted analyses not as a factor of interest but to better assess how covariate measures contribute to understanding of PFT and PRO beyond what might be understood with other available demographic data.

*Image acquisition, processing, and visual assessment*

CXR was performed based on the site's standard protocol and Scadding Stage was determined and provided by the site radiologist. The HRCT was obtained in the supine position without contrast and at full inspiration with the following parameters: 500msec exposure time, standard B35f kernel, 0.75mm thickness, and computed interval of 0.5mm[4]. Images were acquired with a variety of scanners (e-Table 1) after calibration with a phantom and approval by GRADS[5]. CT radiation was adjusted based on body mass index (BMI) and the tube current ranged between 145-374 mA. The peak tube voltage ranged between 100-120 kVp. Detailed summary measures can be found in e-Table 1.

A visual assessment score (VAS) was derived for each HRCT based on the Oberstein score[6] (e-Table 2), denoting the major abnormalities and extent of involvement (none, up to 1/3 of the lung involved, between 1/3 to 2/3 of the lung involved, and more than 2/3s of the lung involved). Additional information was obtained regarding presence of lymphadenopathy, airway and vasculature distortion, presence and distribution (cranial caudal versus axial) of parenchymal opacities, and scored based on Fleischner criteria. The HRCT was electronically transmitted from individual sites to the Genomics Informatics Core (GIC) and a single chest radiologist (CF) blinded to disease diagnosis reviewed all HRCT scans using the above scoring system.

Images were obtained from the GIC in raw DICOM (Digital Imaging and Communications in Medicine) format and were converted to three-dimensional NIfTI (Neuroimaging Informatics Technology Initiative) using dcm2niix (https://github.com/rordenlab/dcm2niix) from the dcm2niir R package. We resampled all scans to 1x1x1mm (or 1 $mm^3$) format and segmented the left and right lungs using segment_lung_lr from the lungct R package[7] (https://github.com/ryansar/lungct); this function uses a combination of thresholding and region-based algorithms for segmentation.

*Decorrelation Filter*

The decorrelation filter measures pairwise correlation to identify features to keep and remove to reduce feature redundancy among retained features and preserve representation of removed features. The filter functions by iteratively selecting a candidate feature to keep according to a ranking function and then discarding all candidate features which are represented by the kept variable according to a prespecified correlation cutoff. Formally, let *A* be the set of variables kept and *D* be the set of variables discarded by the filter. The redundance of a kept variable *x* in *A* and the representation of a discarded variable *x'* in *D* are given, respectively, by:

Redundance of *x* = max of {*y* in *A*-*x*} |Corr(*x,y*)|;

Representation of *x'* =max of {y in *D*}|Corr(x',y)|.

For a specified joint tolerance, the filter guarantees that no retained feature has redundance above tolerance and simultaneously no discarded feature has representation below tolerance. We specified a tolerance of 0.9 on the redundance and representation. We considered two ranking functions. The first is a small set ranking which leads to greedy selection of small kept sets *A* by ranking candidate variables higher if they will result in a larger number of discarded candidates when kept. Specifically, $R(x|k)$ = # {*y* in $C_k$ : |Corr(*x, y*)| ≥ 0.9} with ties broken at random where $C_k$ is the set of candidate variables remaining at iteration *k*. The second is a radiomics-specific modification of the small set ranking in which the rank of second order radiomic features (spatial features) is set to 0 if any first order radiomic features (non-spatial features) remain in the set of candidate variables. A filter with this ranking function selects second order features to keep only when no candidate first order feature remains which may be selected to represent it. This decorrelation filter with first-orderpreferred ranking was used in all analyses to better distinguish contributions of second-order (spatial) radiomic features from first-order (non-spatial) features.

*Validation Analysis*

When applying a decorrelation filter to the radiomics and then clustering, randomness arises in four ways. There is randomness in the sample, in the decorrelation filter, in the initializations of the fitting algorithm, and in the permuted data sets used for estimating the L1 bound and number of clusters. In the following two analyses, we aim to quantify the sensitivity of clustering and secondary analysis results to these four sources of randomness. In the first analysis, we consider simultaneously randomness due to the decorrelation filter, algorithm initialization, and permutations in estimating the number of clusters and L1 bound in RSKC. In the second analysis, using a bootstrap approach, we consider simultaneously randomness due to the sample, the decorrelation filter, and algorithm initialization.

As a first validation analysis, we analyze the radiomics data 512 times using the same approach. First, we apply the decorrelation filter with a small set ranking, which greedily selects small representative sets of radiomic features and breaks ties in rank at random. Using the filtered data, we estimate the number of L1 bound and number of clusters appropriate for clustering using a standardized BCS-based gap statistic from 10 permutations and 10 cluster fitting initializations per setting. Here, the number of clusters is assumed to take a value between 2 and 8 while the L1 bound is assumed to take a value between 1.5 and the square root of the number of features selected by the filter rounded to the nearest 0.5. Using the estimated L1 bound and number of clusters, clusters are fit to the filtered data and cluster labels are recorded.

From these labels, we report the distribution of the pairwise ARI between pairs of the 512 analyses, as well as the maximum ARI between fit clusters and Scadding stage.

Using the labels, we fit two linear models. The first predicts pre-BD FVC from cluster labels and base factors (age, sex, race, BMI, and height) and the second predicts pre-BD FVC from cluster labels, base factors, and Scadding stage. Partial F-test *P*-values are used to quantify significance of cluster label in these models.

We report maximum *P*-values for the cluster and base factor model and for the cluster, Scadding, and base factor model over the 512 analyses, as well as the proportion of *P*-values less than 0.01.

As a second validation analysis, we sample 500 bootstrap data sets from the radiomics data. To each data set, we fit the same number of clusters using the same L1 bound from primary analyses with 10 initializations. Cluster labels and number of unique observations in the data set are recorded. For pairs of analyses, we report the distribution of the ARI of clusters fit to the pair's unique overlapping set of observations as well as the size of the unique overlapping set. We report also the maximum ARI between fit clusters and Scadding stage for each analysis.

Using the labels, we fit two linear models to the bootstrap sample. The first predicts pre-BD FVC from cluster labels and base factors (age, sex, race, BMI, and height) and the second predicts pre-BD FVC from cluster labels, base factors, and Scadding stage. Partial F-test *P*-values are used to quantify significance of cluster label in these models.

We report maximum *P*-values for the cluster and base factor model and for the cluster, Scadding, and base factor model over the 500 analyses, as well as the proportion of *P*-values less than 0.01.

**E-Appendix 2: Results of the Validation Analyses**

In the first validation analysis, pairwise ARI values ranged from 0.3 to 1 and peaked around 0.5 (e-Figure 2). The maximum ARI of fit clusters with Scadding stage was 0.076. In the linear models, in the demographic adjusted and demographic and Scadding stage adjusted model, the maximum *P*-value for the significance of cluster was <0.0001 and the corresponding proportion of *P*-values less than 0.01 was 100%.

In the second validation analysis, bootstrap samples contained between 183 and 218 unique observations and contained 203 on average. Pairs of bootstrap samples contained between 98 and 155 unique overlapping observations and contained 128 on average or about 63% of the sample is used in each ARI calculation. Pairwise ARI values computed from unique overlapping observations ranged from 0.2 to 1 and had a distribution with two peaks around 0.55 and 0.85 (e-Figure 2). The maximum ARI of fit clusters with Scadding stage was 0.17. For significance of cluster label in linear models fit to bootstrap samples, maximum *P*-value for the significant of cluster in the demographic adjusted model was <0.0001 and the proportion of *P*-values less than 0.01 was 100%. In the demographic and Scadding stage adjusted models, the maximum *P*-value for the significance of cluster was 0.02 and the proportion of *P*-values less than 0.01 was 99.8%

**References for Supplementary Material:**


1. HUNNINGHAKE G. Statement on Sarcoidosis. Am J Respir Crit. *Care Med*. 1999;160:736-755.

2. Miller MR, Crapo R, Hankinson J, et al. General considerations for lung function testing. *Eur Respir J*. 2005;26(1):153-161. doi:10.1183/09031936.05.00034505

3. Graham BL, Brusasco V, Burgos F, et al. 2017 ERS/ATS standards for single-breath carbon monoxide uptake in the lung. *Eur Respir J*. 2017;49(1):1600016. doi:10.1183/13993003.00016-2016

4. Zach JA, Newell Jr JD, Schroeder J, et al. Quantitative CT of the lungs and airways in healthy non-smoking adults. *Invest Radiol*. 2012;47(10):596.

5. Moller DR, Koth LL, Maier LA, et al. Rationale and Design of the Genomic Research in Alpha-1 Antitrypsin Deficiency and Sarcoidosis (GRADS) Study. Sarcoidosis Protocol. *Ann Am Thorac Soc*. 2015;12(10):1561-1571. doi:10.1513/AnnalsATS.201503-172OT

6. Oberstein A, von Zitzewitz H, Schweden F, Müller-Quernheim J. Non invasive evaluation of the inflammatory activity in sarcoidosis with high-resolution computed tomography. *Sarcoidosis Vasc Diffuse Lung Dis Off J WASOG*. 1997;14(1):65-72.

7. Ryan SM, Vestal B, Maier LA, Carlson NE, Muschelli J. Template Creation for High-Resolution Computed Tomography Scans of the Lung in R Software. *Acad Radiol*. 2020;27(8):e204-e215. doi:10.1016/j.acra.2019.10.030

**E-Table 1: IBSI Standards Scanner and Scanner Protocol Summary Measures**

| Measure | mean | std | min | max | num.non.na | pct.non.na |
|---|---|---|---|---|---|---|
| KVP | 119.9 | 1.2 | 100 | 120 | 197744 | 1 |
| XRayTubeCurrent | 251.2 | 58.4 | 145 | 374 | 197744 | 1 |
| Exposure | 87.4 | 67.6 | 1 | 190 | 197744 | 1 |
| ExposureTime | 500.9 | 2.5 | 500 | 508 | 197744 | 1 |
| SliceThickness | 0.70 | 0.082 | 0.625 | 3 | 197744 | 1 |
| SpacingBetweenSlices | 0.5 | 0 | 0.5 | 0.5 | 88325 | 0.45 |
| InstanceNumber | 313.0 | 184.0 | 1 | 784 | 197744 | 1 |
| PixelSpacing1 | 0.69 | 0.084 | 0.52 | 0.96 | 197744 | 1 |
| PixelSpacing2 | 0.69 | 0.084 | 0.52 | 0.96 | 197744 | 1 |
| PixelSpacing3 | 0.50 | 0.076 | 0 | 3 | 197744 | 1 |
| SoftwareVersions[a] | | | | | | |
| Manufacturer[b] | | | | | | |
| ManufacturerModelName[c] | | | | | | |
| ConvolutionKernel[d] | | | | | | |

[a]Software Versions: coreload.81, gmp_vct.42, 07MW18.4, gmp_vct.26, sles_hde.84, syngo CT 2012B, syngo CT VA48A, syngo CT 2010A, syngo CT 2009E, 3.2.0, syngo CT 2011A

[b]Manufacturers: GE MEDICAL SYSTEMS, SIEMENS, Philips

[c]Manufacturer Model Names: Optima CT660, LightSpeed VCT, Discovery CT750 HD, SOMATOM Definition Flash, SOMATOM Definition AS+, SOMATOM Definition, Sensation 64, iCT 128

[d]ConvolutionKernel: STANDARD, B35f, B

**E-Table 2:** The distribution of scanner type across radiomic groups to assess the effectiveness of harmonization .

| | | Radiomics Cluster | | | | ***P*-value** |
|---|---|---|---|---|---|---|
| **Scanner Model[a]** | Overall | 1 | 2 | 3 | 4 | >0.9 |
| **Discovery CT750 HD** | 4 (1.3%) | 1 (1.8%) | 2 (1.8%) | 1 (1.9%) | 0 (0%) | |
| **iCT 128** | 34 (11%) | 8 (14%) | 15 (14%) | 3 (5.6%) | 8 (8.0%) | |
| **LightSpeed VCT** | 108 (34%) | 19 (34%) | 38 (35%) | 18 (33%) | 33 (33%) | |
| **Optima CT660** | 3 (0.9%) | 1 (1.8%) | 0 (0%) | 1 (1.9%) | 1 (1.0%) | |
| **Sensation 64** | 8 (2.5%) | 2 (3.6%) | 2 (1.8%) | 1 (1.9%) | 3 (3.0%) | |
| **SOMATOM Definition** | 38 (12%) | 7 (13%) | 13 (12%) | 7 (13%) | 11 (11%) | |
| **SOMATOM Definition AS+** | 91 (28%) | 15 (27%) | 27 (25%) | 17 (31%) | 32 (32%) | |
| **SOMATOM Definition Flash** | 34 (11%) | 3 (5.4%) | 13 (12%) | 6 (11%) | 12 (12%) | |

[a]Discovery = Discovery CT750 HD; iCT = iCT 128; LightSpeed = LightSpeed VCT; Optima = Optima CT660;  Sensation = Sensation 64; SOM Def = SOMATOM Definition; SOM Def AS = SOMATOM Definition AS;  SOM Def Flash = SOMATOM Definition Flash

**E-Table 3: Oberstein component definitions**

| Component | Definition | Scoring (0-3) |
|---|---|---|
| **BVB** | Thickening or irregularity of the bronchovascular bundle | 0=none; 1=1-33% lung volume affected; 2=34-67% lung volume affected; 3=68-100% lung volume affected |
| **PC** | Parenchymal consolidation (including ground-glass opacifications) | 0=none; 1=1-33% lung volume affected; 2=34-67% lung volume affected; 3=68-100% lung volume affected |
| **ND** | Intra-parenchymal nodules | 0=none; 1=1-33% lung volume affected; 2=34-67% lung volume affected; 3=68-100% lung volume affected |
| **LS** | Septal and nonseptal lines | 0=none; 1=1-33% lung volume affected; 2=34-67% lung volume affected; 3=68-100% lung volume affected |
| **PLT** | Focal pleural thickening | 0=none; 1=mild; 2=moderate; 3=severe |
| **LN** | Enlargement of the lymph nodes (short axis >1cm) | 0=none; 1=mild; 2=moderate; 3=severe |

**e-Table 4: Oberstein components by radiomic cluster and differences in distributions compared using simulated Fisher's *P*-values to address small cell sizes.**

| | Overall | 1 | 2 | 3 | 4 | |
|---|---|---|---|---|---|---|
| | N=317 | N=56 | N=108 | N=53 | N=100 | *P*-value |
| **Oberstein_BVB** | | | | | | < 0.01 |
| **0** | 100 (31.5%) | 28 (50.0%) | 53 (49.1%) | 8 (15.1%) | 11 (11.0%) | |
| **1** | 79 (24.9%) | 20 (35.7%) | 29 (26.9%) | 13 (24.5%) | 17 (17.0%) | |
| **2** | 72 (22.7%) | 5 (8.9%) | 21 (19.4%) | 15 (28.3%) | 31 (31.0%) | |
| **3** | 66 (20.8%) | 3 (5.4%) | 5 (4.6%) | 17 (32.1%) | 41 (41.0%) | |
| **Oberstein_PC** | | | | | | < 0.01 |
| **0** | 155 (48.9%) | 46 (82.1%) | 74 (68.5%) | 19 (35.8%) | 16 (16.0%) | |
| **1** | 63 (19.9%) | 6 (10.7%) | 20 (18.5%) | 15 (28.3%) | 22 (22.0%) | |
| **2** | 65 (20.5%) | 4 (7.1%) | 14 (13.0%) | 11 (20.8%) | 36 (36.0%) | |
| **3** | 34 (10.7%) | 0 (0.0%) | 0 (0.0%) | 8 (15.1%) | 26 (26.0%) | |
| **Oberstein_ND** | | | | | | < 0.01 |
| **0** | 147 (46.4%) | 35 (62.5%) | 57 (52.8%) | 18 (34.0%) | 37 (37.0%) | |
| **1** | 109 (34.4%) | 14 (25.0%) | 39 (36.1%) | 18 (34.0%) | 38 (38.0%) | |
| **2** | 38 (12.0%) | 3 (5.4%) | 9 (8.3%) | 11 (20.8%) | 15 (15.0%) | |
| **3** | 23 (7.3%) | 4 (7.1%) | 3 (2.8%) | 6 (11.3%) | 10 (10.0%) | |
| **Oberstein_LS** | | | | | | < 0.01 |
| **0** | 232 (73.2%) | 51 (91.1%) | 85 (78.7%) | 35 (66.0%) | 61 (61.0%) | |
| **1** | 82 (25.9%) | 5 (8.9%) | 23 (21.3%) | 17 (32.1%) | 37 (37.0%) | |
| **2** | 3 (0.9%) | 0 (0.0%) | 0 (0.0%) | 1 (1.9%) | 2 (2.0%) | |
| **Oberstein_PLT** | | | | | | 0.35 |
| **0** | 311 (98.1%) | 56 (100.0%) | 107 (99.1%) | 51 (96.2%) | 97 (97.0%) | |

| | | | | | | |
|---|---|---|---|---|---|---|
| **1** | 5 (1.6%) | 0 (0.0%) | 1 (0.9%) | 1 (1.9%) | 3 (3.0%) | |
| **2** | 1 (0.3%) | 0 (0.0%) | 0 (0.0%) | 1 (1.9%) | 0 (0.0%) | |
| **Oberstein_LN** | | | | | | 0.01 |
| **0** | 136 (42.9%) | 35 (62.5%) | 47 (43.5%) | 20 (37.7%) | 34 (34.0%) | |
| **1** | 88 (27.8%) | 14 (25.0%) | 29 (26.9%) | 19 (35.8%) | 26 (26.0%) | |
| **2** | 90 (28.4%) | 7 (12.5%) | 30 (27.8%) | 14 (26.4%) | 39 (39.0%) | |
| **3** | 3 (0.9%) | 0 (0.0%) | 2 (1.9%) | 0 (0.0%) | 1 (1.0%) | |

**e-Table 5:** Results of the regression analysis of the five discriminatory radiomic measures for PRO's. Each linear regression model included all five radiomic features and was additionally adjusted for age, sex, race, BMI and height. Bolded cells are statistically significant (p-values in the footnote). GLCM - Gaussian is the sum of the GLCM with a Gaussian weight applied. GLCM-Inv Gaussian is the sum of the GLCM for the inverse Gaussian weighting scheme. GLCM Sum Entropy is a measure of the disorder of the GLCM. GLCM Min is the minimum of the GLCM.

| PRO | GLCM Gaussian | GLCM Inv Gaussian | Kurtosis | GLCM Sum Energy | GLCM Min | *P*-value[b] | Rsq | Rsq - base |
|---|---|---|---|---|---|---|---|---|
| **FAS** | 1.50 (1.21) | 0.11 (0.88) | 1.00 (0.90) | 0.85 (1.00) | 0.54 (0.96) | 0.151 | 0.112 | 0.069 |
| **GERDQ** | -0.12 (0.35) | 0.35 (0.27) | -0.40 (0.25) | 0.08 (0.29) | -0.19 (0.28) | 0.201 | 0.063 | 0.04 |
| **CFQ** | 3.32 (2.86) | 0.56 (2.20) | -0.73 (2.09) | 4.02 (2.35) | 0.69 (2.29) | 0.465 | 0.067 | 0.053 |
| **SOBQ** | 4.24 (3.69) | 3.61 (2.82) | -0.05 (2.69) | -4.62 (3.03) | -0.61 (2.96) | <0.0001 | 0.18 | 0.111 |
| **Promis** | **3.37 (1.56)**[a] | 2.34 (1.19) | 1.08 (1.14) | 0.94 (1.30) | -1.32 (1.27) | 0.258 | 0.054 | 0.031 |
| **SF12 - Physical** | 0.75 (1.81) | -0.25 (1.39) | 0.64 (1.32) | 2.22 (1.49) | 0.74 (1.45) | 0.006 | 0.155 | 0.109 |
| **SF12 - Mental** | -0.94 (1.69) | -1.48 (1.30) | 0.72 (1.24) | -0.04 (1.40) | -0.75 (1.36) | 0.706 | 0.066 | 0.057 |

[a] P-value <0.0032; [b] Overall P-value testing whether any of the radiomic measures are significant in the regression model.

**E Figure 1:** Diagram of patient population leading to analysis datasets (N=321 and N=318).

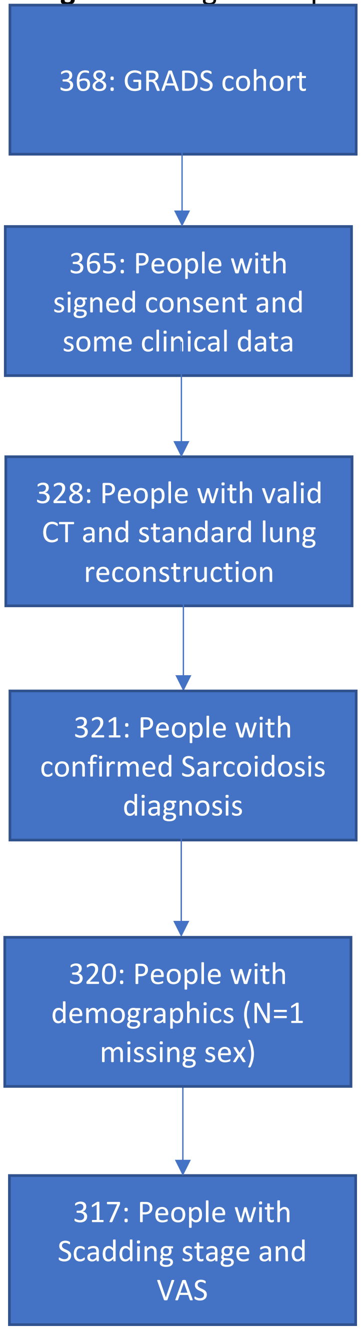

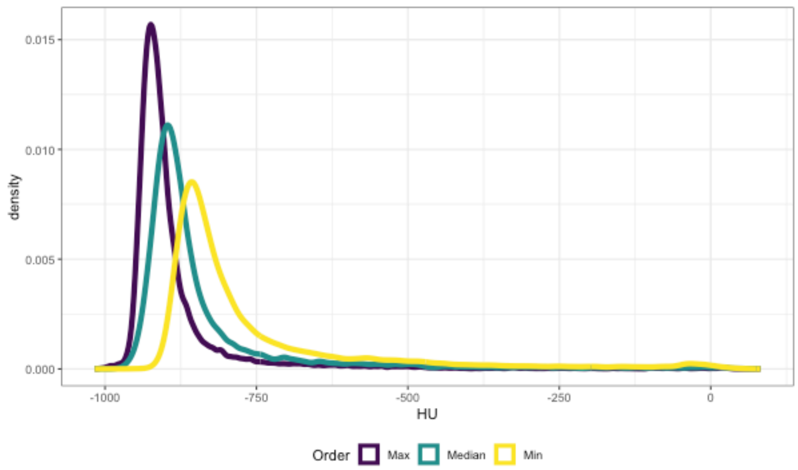


**E-Figure 2:** Distribution of HU from HRCT images with the maximum, median, and minimum kurtosis.

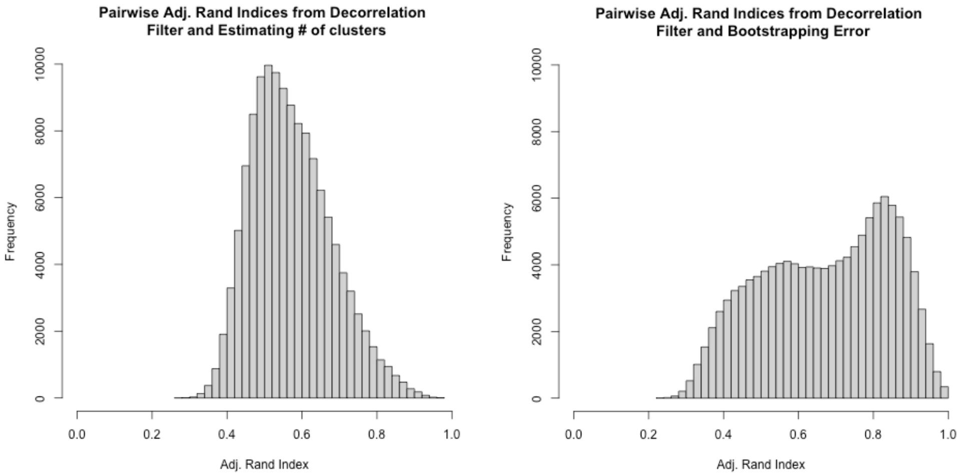


**E-Figure 3:** Results from the validation studies. Left shows the distribution of ARI for scenario 1. Right shows the distribution of ARI for scenario 2.
